# Impact of exposure frequency on disease burden of the common cold - a mathematical modeling perspective

**DOI:** 10.1101/2024.04.26.24306416

**Authors:** Sebastian Gerdes, Michael Rank, Ingmar Glauche, Ingo Roeder

## Abstract

The common cold is a frequent disease in humans and can be caused by a multitude of different viruses. Despite its typically mild nature, the high prevalence of the common cold causes significant human suffering and economic costs. Oftentimes, strategies to reduce contacts are used in order to prevent infection. To better understand the dynamics of this ubiquitous ailment, we develop two novel mathematical models: the common cold ordinary differential equation (CC-ODE) model at the population level, and the common cold individual-based (CC-IB) model at the individual level. Our study aims to investigate whether the frequency of population / individual exposure to an exemplary common cold pathogen influences the average disease burden associated with this virus.

On the one hand, the CC-ODE model captures the dynamics of the common cold within a population, considering factors such as infectivity and contact rates, as well as development of specific immunity in the population. On the other hand, the CC-IB model provides a granular perspective by simulating individual-level interactions and infection dynamics, incorporating heterogeneity in contact rates.

By employing these models, we explore the impact of exposure frequencies upon the net disease burden of common cold infections in theoretical settings. In both modeling approaches, we show that under specific parameter configurations (i.e., characteristics of the virus and the population), increased exposure can result in a lower average disease burden. While increasing contact rates may be ethically justifiable for low-mortality common cold pathogens, we explicitly do not advocate for such measures in severe illnesses. The mathematical approaches we introduce are simple yet powerful and can be taken as a starting point for the investigation of specific common cold pathogens and scenarios.

## 1 INTRODUCTION

### 1.1 Biological background

The common cold is a frequent disease in humans and is generally caused by viral infection of the upper respiratory tract (Thomas and Bomar, 2022). Although mild in most cases, it poses a significant disease burden on individuals and societies, both in terms of human suffering and economic loss. The common cold is the most frequent illness in the US with approximately 25 million documented cases per year (Passioti et al., 2014). It is estimated that in the US alone, the economic cost of the common cold is approximately $25 billion per year (Bramley et al., 2002). Despite the large number of cases and the great associated disease and economic burden the common cold is currently not a prioritized research topic. A thorough and precise understanding of the disease dynamics both an a societal and an individual level are lacking today, but might pave the way to better prevention and treatment strategies.

The most frequent causative viral agents are rhino viruses (approx. 30% to 50%), corona viruses (approx. 10% to 15%, not including SARS-CoV-1, SARS-CoV-2 and MERS), influenza viruses (approx. 5% to 15%), respiratory syncytial viruses (approx. 5%), parainfluenza viruses (approx. 5%), adeno viruses (less than 5%), entero viruses (less than 5%), and further unknown viruses (approx. 20% to 30%) (Heikkinen and Järvinen, 2003). Of these viruses, different strains are circulating and they are constantly subjected to genetic shift and drift.

The immune system is generally capable of clearing a common cold without additional treatment. In healthy individuals the symptoms are often relatively mild (including sneezing, stuffy nose, runny nose, sore throat, coughing, post-nasal drip, watery eyes, fever). Most often, the intensity of symptoms peaks around day 3 or 4 and around day 7 recovery begins (Heikkinen and Järvinen, 2003). The median duration of symptoms of a common cold has been estimated to be approximately 11 days (Arruda et al., 1997).

In fighting a common cold, different components of the immune system are involved (Murphy et al., 2022). Generally, the infectious agent enters via the mucous membranes. Here, both specific and unspecific components of the immune system can often already eliminate the infectious agent. In this case, the exposure may result in an asymptomatic course, however, possibly involving training of the immune system. If the infectious agent settles and proliferates, typical symptoms of a common cold may develop. Over time, more powerful components of the specific immune system come into play. In particular, the specific immune system continuously improves its ability to recognize the pathogen and efficiently eliminates it.

After the infection has subsided, the immune system usually retains the ability to recognize the respective pathogen for a while. Hence, after immediate reexposure, it is unlikely that another symptomatic infection occurs. However, with time the newly acquired specific immunity generally deteriorates and may even revert to the baseline level. Furthermore, it is important to note that there is significant cross-reactivity between different virus strains, e.g. in the case of rhinoviruses (Glanville et al., 2013), and possibly even between different viruses. Therefore, a symptomatic infection may be alleviated or prevented after exposure to a virus, if an infection with a similar virus or virus strain has occurred previously (see figure 1).

**Figure 1.**
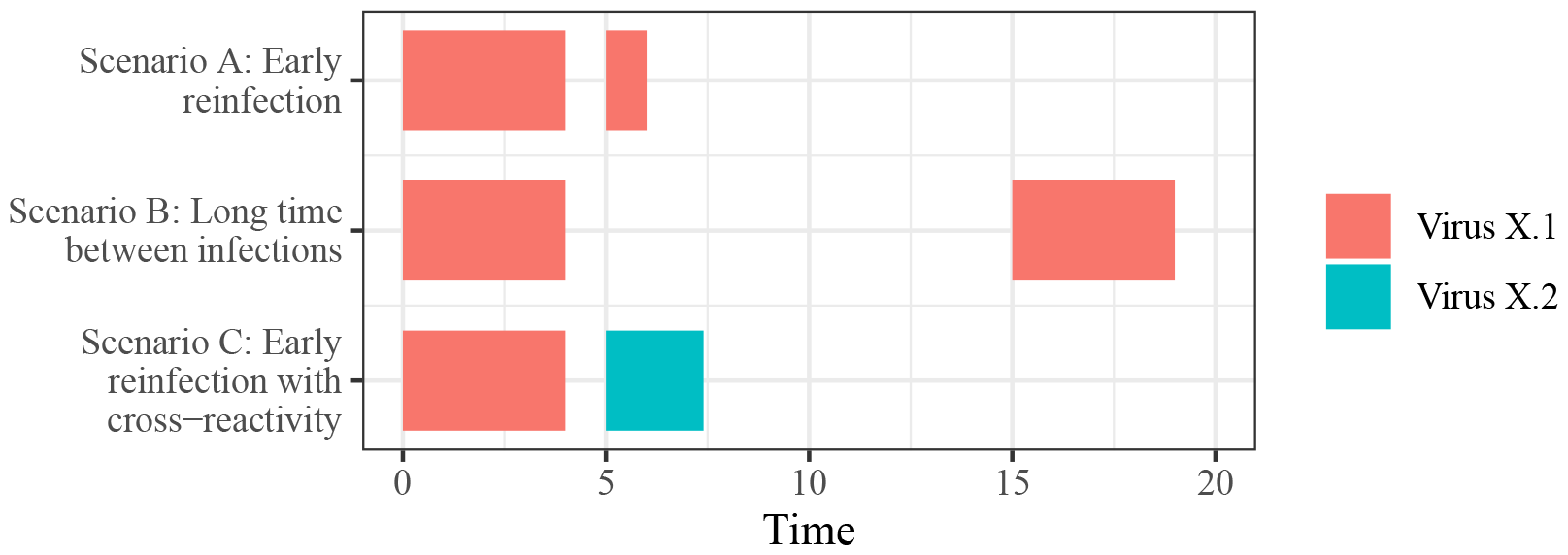
Schematic representation of different hypothetical scenarios for immunity upon reinfection with an exemplary virus denoted by “X” on a time scale without units. In scenario A, upon immediate reinfection with strain “X.1” of the exemplary virus “X”, symptoms are likely to be mild and the infection is usually short or the infection might even be asymptomatic. In scenario B the time between infections is long, and immunity will likely be lost for many viruses and duration of the infection would be long again and might be associated with more severe symptoms. Scenario C considers reinfection with an exemplary strain “X.2” of virus X. In this case there might be some cross-reactivity, which alleviates symptoms in case of infection with the related virus strain “X.2”.

The degree of persistence of specific immunity seems to differ considerably between different common cold pathogens (Turner, 2015). Infections with rhinoviruses and adenoviruses seem to generally result in long-term, protective immunity against the specific virus serotype, however, not against other serotypes of the virus. Infections with coronaviruses, parainfluenzaviruses, RS-viruses and multiple other common-cold-viruses seem to usually result in short-term immunity, that declines over time.

There is a large corpus of mathematical models describing the dynamics of infectious diseases in a population. Among these models, the classical Susceptible-Infected-Recovered (SIR) epidemic model introduced by (Kermack and McKendrick, 1932) holds a prominent place. It is an ordinary differential equation (ODE) model, in which a susceptible fraction of the population (compartment **S**) can get infected with an infectious disease and hence transition to compartment **I**. Finally, individuals in compartment **I** recover and hence pass into compartment **R**, in which they are immune to the infectious disease. There are numerous variants of this model. One variant that is relevant in the context of this paper is the SIRS model, in which immunity in compartment **R** is lost with a certain rate and individuals hence transition from compartment **R** back into compartment **S**. Another variant of the SIR model is the SIS model (first described by Weiss and Dishon, 1971), in which no immunity is acquired (no compartment **R**) and individuals pass directly into compartment **S** when recovering.

### 1.2 Objectives

The relation between disease burden per capita per time (referred to as *mean/average disease burden* from hereon) and the exposure frequency to particular viruses typically causative of common colds has not been explicitly studied. Exposure frequency may depend among other aspects on overall interpersonal contact patterns and contact reduction strategies of infected individuals. In a certain range, increased exposure likely increases the spread of viral disease and, thus, increases the mean disease burden for most viruses causing common colds. However, we want to investigate if this relation might reverse under certain circumstances, if a critical exposure frequency is exceeded due to increased training of the immune system. In other words, more contacts and hence more exposure to pathogen might strengthen the immune system so that for some pathogens the disease burden related to this virus might be reduced. Even though this seems plausible, there are neither reliable epidemiological data available demonstrating such an effect for the common cold, nor has this aspect been studied explicitly in a mathematical modeling framework to our knowledge.

Therefore, we aim to investigate this hypothesis by applying a mathematical modeling approach. Mathematical models express hypotheses in formal, quantitative terms and can be used to evaluate the implications of different hypotheses and design informative experiments. In the following, we present two novel, related mathematical models in order to address the aforementioned research question.

In our analysis, we consider an exemplary virus that can cause an upper respiratory infection. The proposed methods can be applied to any real common cold pathogen by choosing appropriate parameter configurations or estimating them from respective data. Our goal is to find a simple and universal mathematical framework that describes the progression of the common cold rather than the precise analysis of one specific pathogen.

The first model that we call **CC-ODE model** (*common cold ordinary differential equation model*) is an ODE model based on the SIRS model. A formal description of the model can be found in section 2.1 and analysis results in section 3.1. The second model is an individual-based model derived from the SIS model and is referred to as **CC-IB model** (*common cold individual-based model*). It is described in section 2.2 and section 3.2. Individual-based models allow to follow individuals and their properties (DeAngelis and Grimm, 2014). Thereby, they are capable of representing stochasticity and autonomy of individuals. Complex, non-linear phenomena may emerge as a result of individuals that follow simple rules.

Our work is based on the prototypical SIR model family, which allows for an easy connection and integration into the current scientific discourse. In case of the ODE approach, the plain SIR model cannot account for loss of immunity, which is essential to the goals of the work presented in this paper. Therefore, we choose the slightly more complex SIRS model, in which immunity can be lost. In case of the individual-based approach, complex phenomena can be represented by individual states of immunity. Hence, the explicit modeling of the **R** compartment is not necessary and the SIS model is sufficient as a basis. The two chosen modeling approaches shall complement each other, the CC-ODE model being better suited for deriving analytical results in closed form and deriving population-centered results, while the CC-IB model inherently allows to represent heterogeneity among individuals, stochasticity and tracking of individual fates.

## 2 METHODS

### 2.1 CC-ODE model

In this section, the **CC-ODE model** (*common cold ordinary differential equation model*) is presented and described. It is an ODE model based on the SIRS model, a classical model describing the dynamics of an infectious disease on a population level. In the SIRS model, individuals can switch between three different compartments. First, the susceptible individuals are in compartment **S**. They do not carry the disease and can potentially get infected. The number of individuals in this compartment at time *t* is given by *S* = *S*(*t*). Second, the infected individuals are in compartment **I** and can spread the disease. The number of individuals in this compartment at time *t* is denoted by *I* = *I*(*t*). Third, the recovered (and immune) individuals are in compartment **R**. They can neither acquire nor spread the disease and the number of individuals at time *t* reads *R* = *R*(*t*). The total number of individuals *N* (*t*) = *S* + *I* + *R* is fixed (phenomena such as birth, death and migration are not included in the model). For reasons of simplicity, we assume *N* = 1 so that *S, I* and *R* can be interpreted as proportions of a population that is constant in size. The model equations read

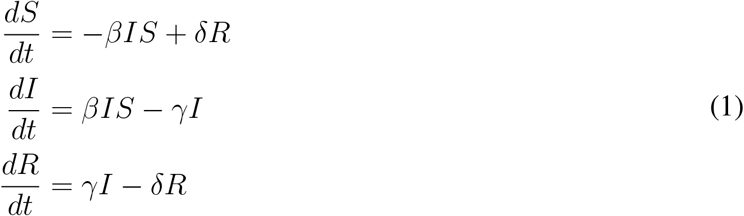

with initial conditions *S*(0), *I*(0) and *R*(0). The model parameters are

- *β* ≥ 0: infection rate (influenced by contact rate and infectivity of the pathogen),
- *γ* ≥ 0: recovery rate,
- *δ* ≥ 0: immunity loss rate.

Note that by setting *δ* = 0 we obtain the basic SIR model. This model describes the dynamics of a population exposed to one single virus without interaction to other pathogens or concurrent virus strains. An infection with this virus (strain) can lead to immunity that eventually subsides, which holds true for most common cold pathogens.

To investigate if a higher contact rate can eventually lead to a reduced overall disease burden, we introduce some amendments of the equations and parameters leading to an ODE model, which we call *CC-ODE model*. We assume that an increase of the contact rate leads to shorter (or less intense) infections due to development of specific immunity. Hence, the higher the contact rate, the higher also the recovery rate. However, there is no evidence that the infectivity of the pathogen should also affect the recovery rate. This is why we split the infection rate *β* into two independent factors that describe the infection rate by a contact rate (*β*_2_) and a measure of infectivity (*β*_1_), which can be interpreted as the probability of infection upon exposure:

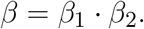

In order to represent the possibility of development of specific immunity to the pathogen (= habituation effect), we introduce the parameter *α* representing the immunogenicity of the virus, giving rise to an additional term *αβ*_2_*I* in the model equations:

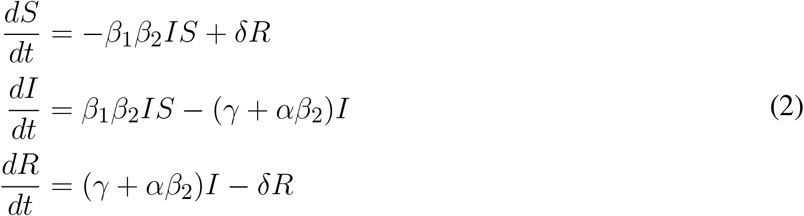

with the additional model parameters

- *β*_1_ ≥ 0: infectivity,
- *β*_2_ ≥ 0: contact rate,
- *α* ≥ 0: immunogenicity, i. e. a measure of the degree of development of specific immunity to the pathogen in the population upon exposure.

*β*_1_ depicts the infectivity of the pathogen. Large values of *β*_1_ correspond to highly contagious diseases. The contact rate *β*_2_ describes the number of contacts per individual in society, with high values indicating a very active population with many contacts. In this way, the higher both of these parameters, the higher the total infection rate. If *β*_1_ = 0 (non-contagious disease) or *β*_2_ = 0 (absolute isolation), there is no spread of the disease. The parameter *α* describes the immunogenicity of the virus, i. e. the immunological habituation effect of the specific immune system against the virus in question. For *α* = 0, there is no development of specific immunity at all and we obtain the classical SIRS model. For *α* + and *β*_2_ *>* 0, development of specific immunity is so effective that infections are eliminated immediately.

This model is applied to describe the dynamics of the common cold on a *population level*. We aim to investigate the disease dynamics in the CC-ODE model by introducing an infection into a small proportion of a population without prior immunity . Thus, the initial conditions read *S*(0) = 0.99, *I*(0) = 0.01 and *R*(0) = 0 for all our simulations of the CC-ODE model. In the following section, we present a novel agent-based model focusing on the *individual level*.

### 2.2 CC-IB model

The **CC-IB model** (*common cold individual-based model*) is derived from the SIS model, in which the compartment with recovered individuals is omitted. In order to facilitate a translation into a corresponding individual-based model, the number of individuals *N* is not set to 1 as in the previous section. Instead, we are dealing with a fixed number *N >* 1 of individuals. The dynamics of the SIS model are described by the following equations:

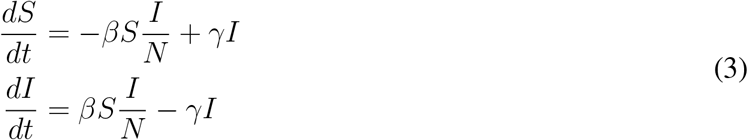

As in the SIR model, the parameter *β* determines infection rate (composed of number of contacts per person per time and the contagion infectivity), while the parameter *γ* determines the recovery rate.

The CC-IB model is derived from this model by assuming a population of *N* individuals, which can switch between **S** and **I** according to probabilistic rules. Each individual can have an individual value for the infection rate *β*, the value of the *i*-th individual denoted *β*_*i*_ (*i* = 1, 2, 3, …, *N*). Since the infectivity of a virus is an inherent, non-changing property of a specific virus, modulations of *β*_*i*_ directly represent changes in the contact rate in the model. The model operates on a discrete time-scale. Each timestep, an individual in **S** acquires an infection (and hence switches to **I**) with probability *β*_*i*_*I/N* :

A diseased individual currently residing in **I** recovers (and hence switches to **S**) with a probability that is given by a transition function *f* (*θ*_*i*_) per timestep, where *θ*_*i*_ denotes the time since the last recovery of the *i*-th individual. *f* (*θ*_*i*_) is defined as the sum of a term representing the ability of the untrained immune system to clear an infection (*c*) and a term representing immunity due to previous exposure to the virus. The term representing specific immunity decays exponentially with rate *dθ*_*i*_, where *d* defines the timescale of the immunological memory:

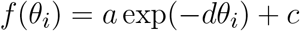

Hence, in summary for each individual identified by index *i*, the probabilities to switch from one state to the other read as follows:

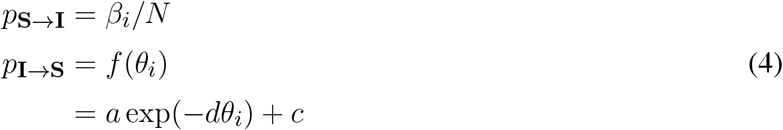

In the simulations, three different parameterizations of *f* (*θ*_*i*_), i. e. three different sets of values for *a, d* and *c*, are considered, corresponding to scenarios denoted “No specific immunity” (*a* = 0, *d* = 0.01, *c* = 0), “Medium specific immunity” (*a* = 0.49, *d* = 0.1, *c* = 0.01) and “Strong specific immunity” (*a* = 0.99, *d* = 0.05, *c* = 0.01). The resulting transition functions are visualized in figure 2.

**Figure 2.**
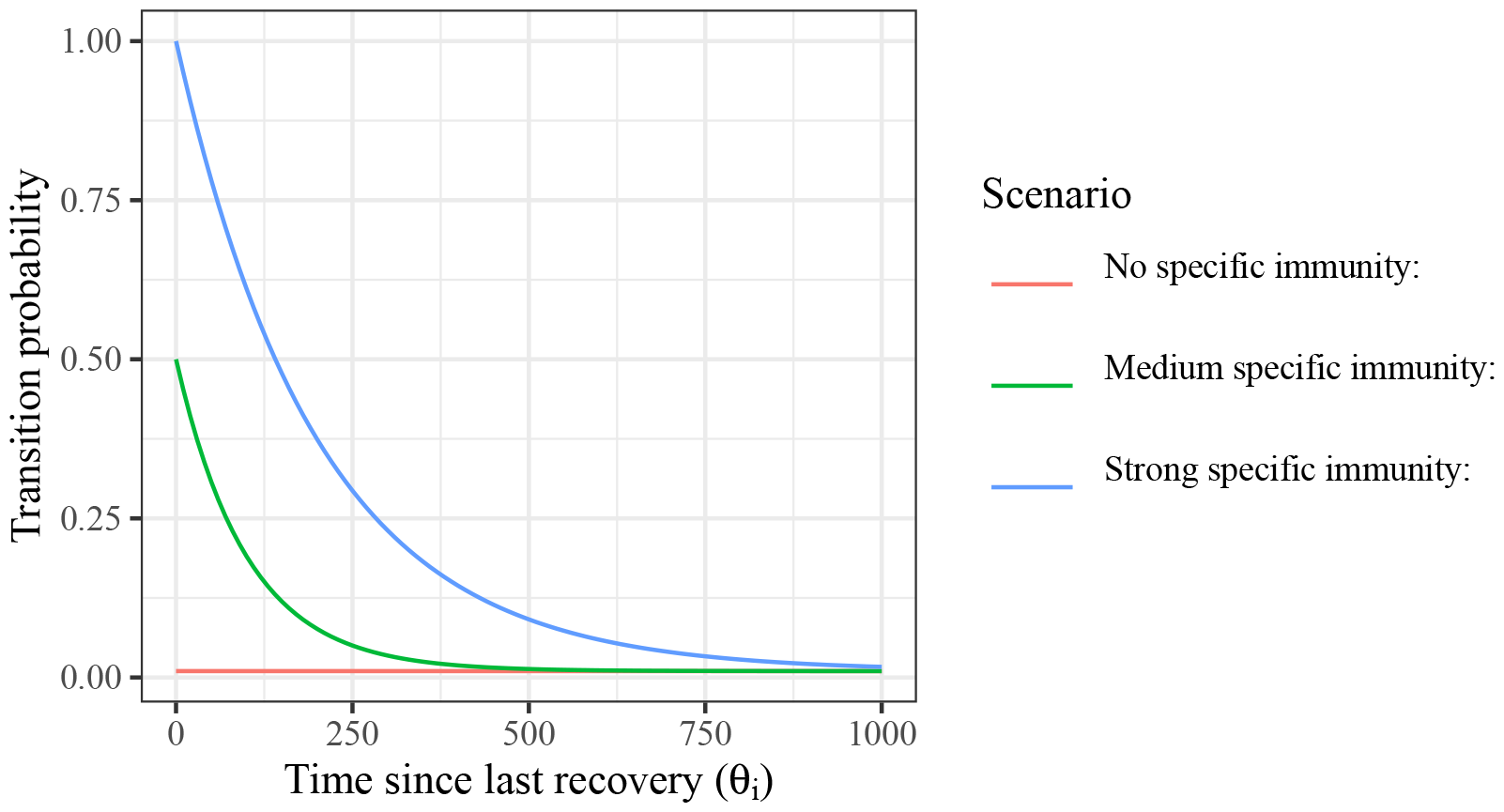
Transition function for the chosen scenarios that are analyzed subsequently. In the first scenario, no training of the immune system is assumed (red line). In the second scenario, it is assumed that an infection results in a mild immunity, which is decaying with time (green line). In the third scenario, it is assumed that infection results in a strong immunity, which, however, is also decaying with time (blue line).

The CC-IB model is simulated 10,000 timesteps via Monte Carlo simulations. The population is comprised of 50 “test individuals”, whose trajectory is followed individually in graphical representations, and 950 additional individuals ensuring a sufficient population size. In the test individuals, *β*_*i*_ ranges from 0.001 to 0.5 equally spaced on a log scale. In the additional individuals, *β*_*i*_ is sampled from a log-normal distribution with *µ* = -4 and *σ* = 1. Initially, without loss of generality, 10 randomly chosen individuals are infected (*I*(0) = 10), the other individuals are susceptible (*S*(0) = 990). It is assumed that none of the individuals have been exposed to the virus before (*θ*_*i*_ = ∞ for all individuals initially).

A graphical overview of the two presented models and their underlying variants can be found in figure 3.

**Figure 3.**
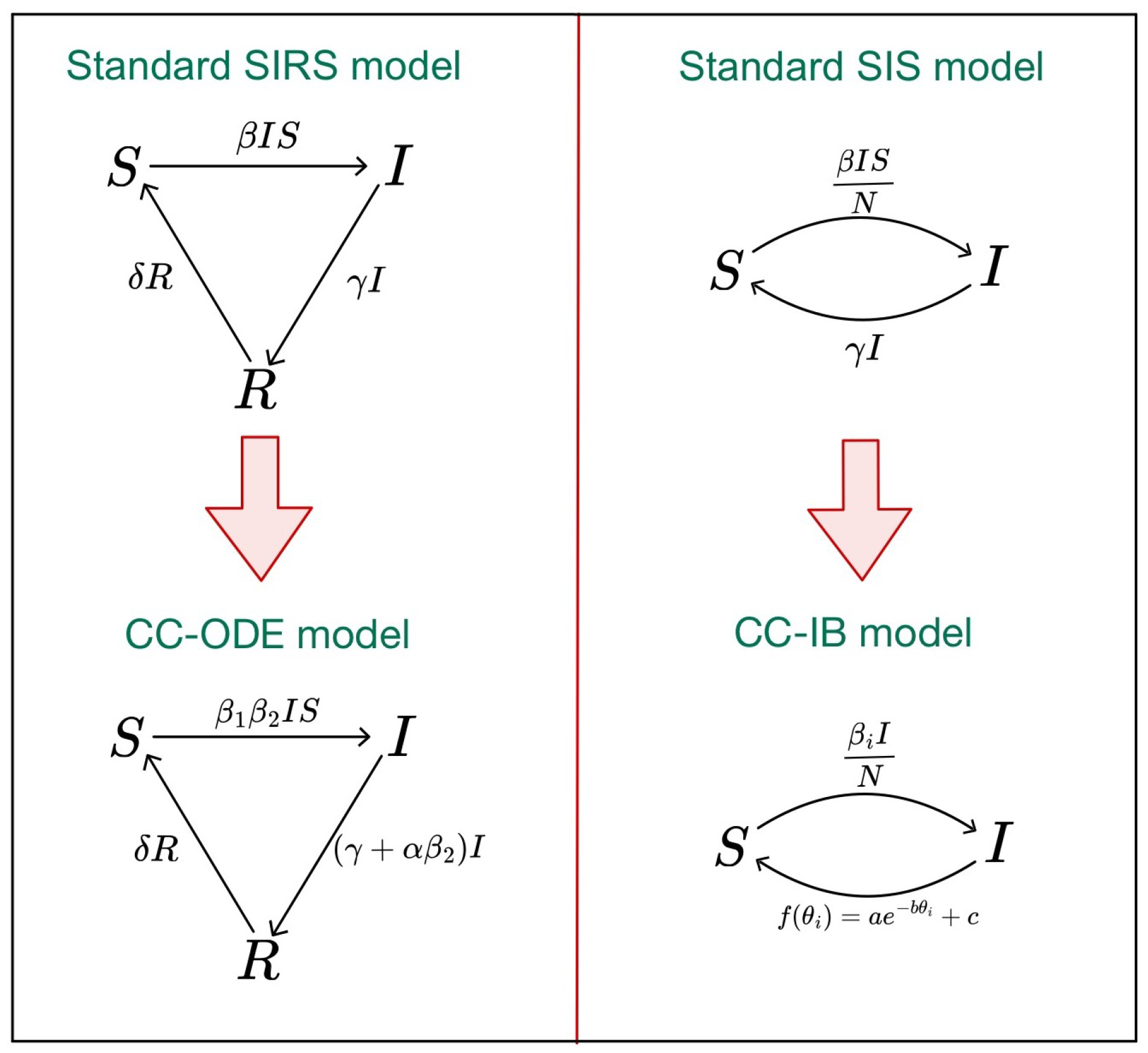
Overview of the underlying standard SIRS model (upper left corner) and standard SIS model (upper right corner), as well as the novel developed CC-ODE model (lower left corner) and CC-IB model (lower right corner). The complete model equations can be found in equation 1 (SIRS model), equation 2 (CC-ODE model), equation 3 (SIS model) and equation 4 (CC-IB model).

## 3 RESULTS

### 3.1 CC-ODE model

The model dynamics for some parameter choices of *α* and *β*_2_ are shown in figure 4. The parameters *α* and *β*_2_ are of primary interest in this study and are varied systematically in the following. The parameters *β*_1_, *γ* and *δ* are held constant (*β*_1_ = 0.7, *γ* = 0.1 and *δ* = 0.1) in the presented analysis in order to reduce complexity, but could in principle be estimated from biological data, in case it is available for specific viruses.

**Figure 4.**
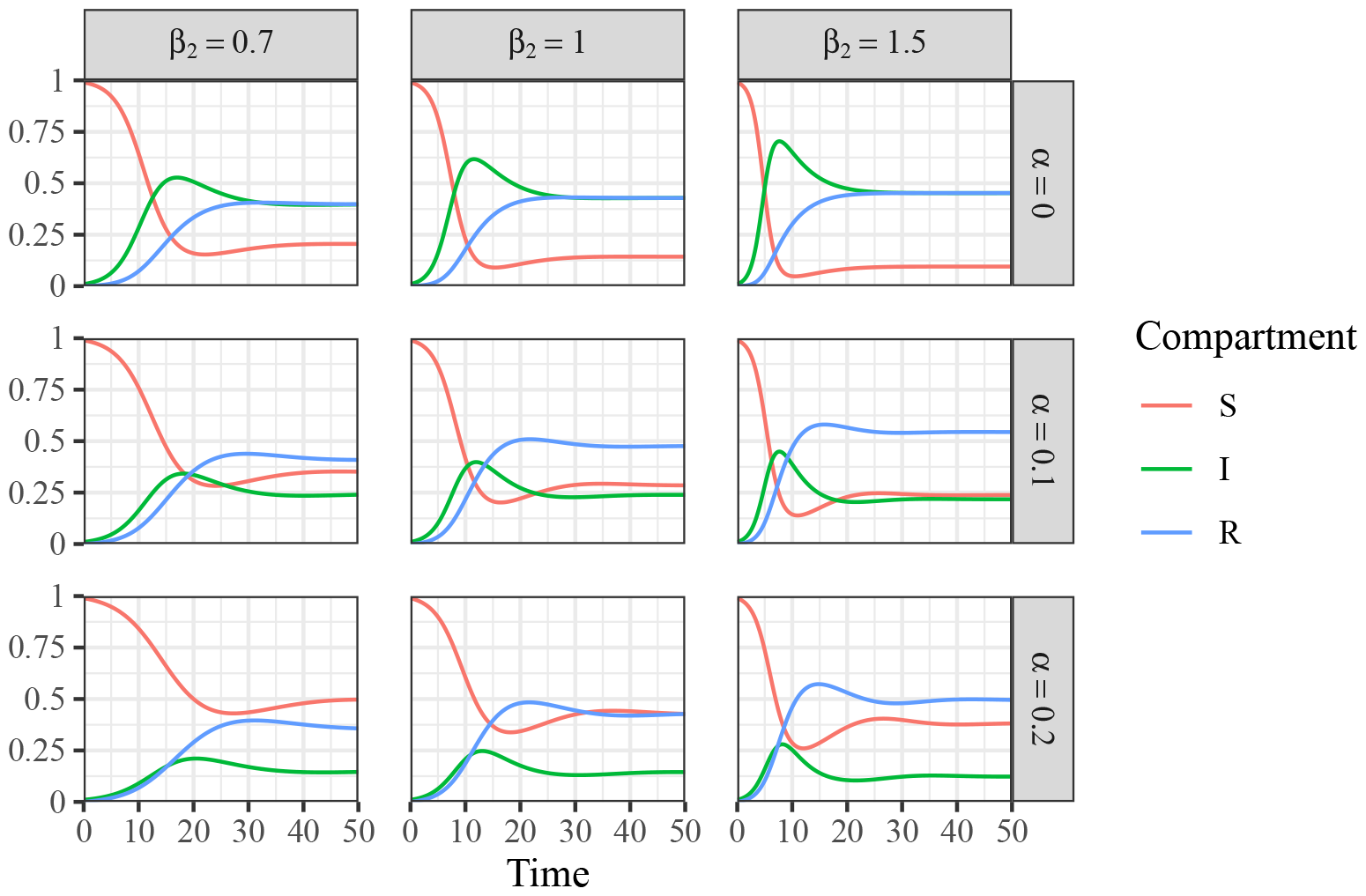
Solutions of CC-ODE model for *t ∈* [0, 50] with initial conditions *S*(0) = 0.99, *I*(0) = 0.01, *R*(0) = 0, parameter values *β*_1_ = 0.7, *γ* = *δ* = 0.1 and different choices for *α ∈ {*0, 0.1, 0.2*}* and *β*_2_ *∈ {*0.7, 1.0, 1.5*}*.

As one would expect, with an increasing value of *α* and constant values for all other parameters, there are more susceptible and less infected individuals. The dynamics for varying *β*_2_ appear to be a bit more complex and depend essentially on the choice of *α*. To further investigate this, we have a closer look on the steady states of the ODE system. There are two steady states of the ODE system. One of them is the trivial steady state (*S*^*\**^, *I*^*\**^, *R*^*\**^) = (1, 0, 0), in which there are solely susceptible individuals in **S**, while there are neither infected nor immune individuals in the population. Hence, there is no infection at all that could be spread and the entire population remains susceptible and healthy. The condition

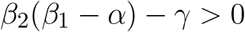

ensures instability of the trivial steady state (*S*^*\**^, *I*^*\**^, *R*^*\**^) = (1, 0, 0) and hence the basic reproduction number reads

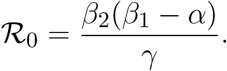

As a consequence, there is a disease outbreak, if ℛ_0_ *>* 1. Otherwise, an introduction of the virus to a small part of the population leads to the extinction of the disease. On the one hand, the higher the probability of infection upon exposure *β*_1_ and contact rate *β*_2_, the higher the chance that the disease is breaking out. On the other hand, the greater the immunogenicity of the virus *α* and the recovery rate *γ*, the lower the the probability that the disease is breaking out. This coincides well with the intuitive notion of these model parameters.

Additionally, there is another steady state, which is non-trivial:

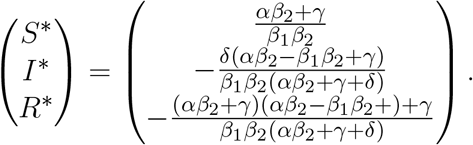

In the following, we focus on this second steady state. Thereby, we further investigate *I*^*\**^ and its behavior depending on the parameters *α* and *β*_2_, see figure 5 a). *I*^*\**^ can be interpreted as the proportion of infected individuals in the long term and is therefore a suitable representation of the mean disease burden after an initial period. For *α >* 0, we obtain curves with one single maximum at

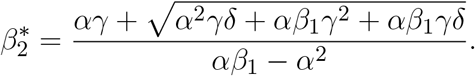

Hence, for 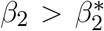, there is a decrease of *I*^*\**^ and thus of the number of infections in the long term. Depending on the specific characteristics of a virus and the resulting different parameters in the ODE model, a higher contact rate of the individuals can lead to an overall lower disease burden. That is, having overall more contacts helps to reduce the average disease burden, if there is sufficient development of specific immunity. Interestingly, with increasing probability of infection upon exposure *β*_1_ the location of the maximum 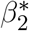 is decreasing, see figure 5 b). This means that for pathogens with higher probability of infection upon exposure, it might be the better strategy to have also a higher contact rate to keep the average disease burden lower. Note, however, that this may not be reasonable for all diseases. The stability analysis of the steady states can be found in the supplementary material.

**Figure 5.**
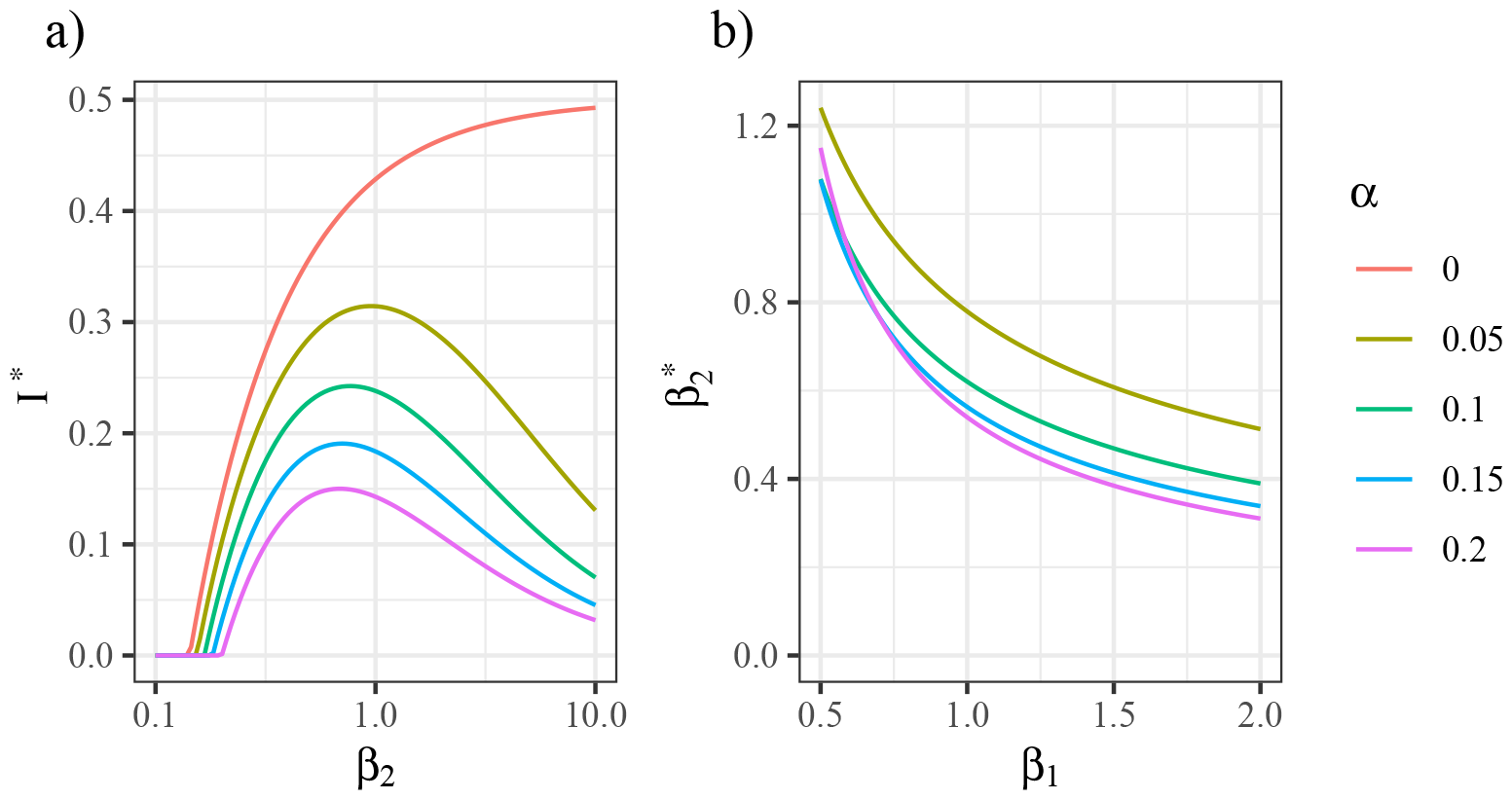
a) Analytical solutions of steady state component *I*^*\**^ (infected individuals) vs. log-scaled contact rate *β*_2_ and *β*_1_ = 0.7. b) Maximum location of steady state 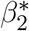 vs. pathogen infectivity *β*_1_. The colors code for different choices of the immunogenicity *α*. Other parameter values are *γ* = *δ* = 0.1.

In order to shed more light on the dynamics at the individual level, we investigate the behavior of the individual-based model (CC-IB model) in the following.

### 3.2 CC-IB model

For the CC-IB model, we evaluate three scenarios corresponding to different patterns of immunity development (no specific immunity, medium specific immunity, strong specific immunity). The number of persons in the compartments **S** and **I** over time is depicted in figure 6. It can be seen that in all three scenarios, the fraction of infected individuals (*I/N*) oscillates around an equilibrium point.

**Figure 6.**
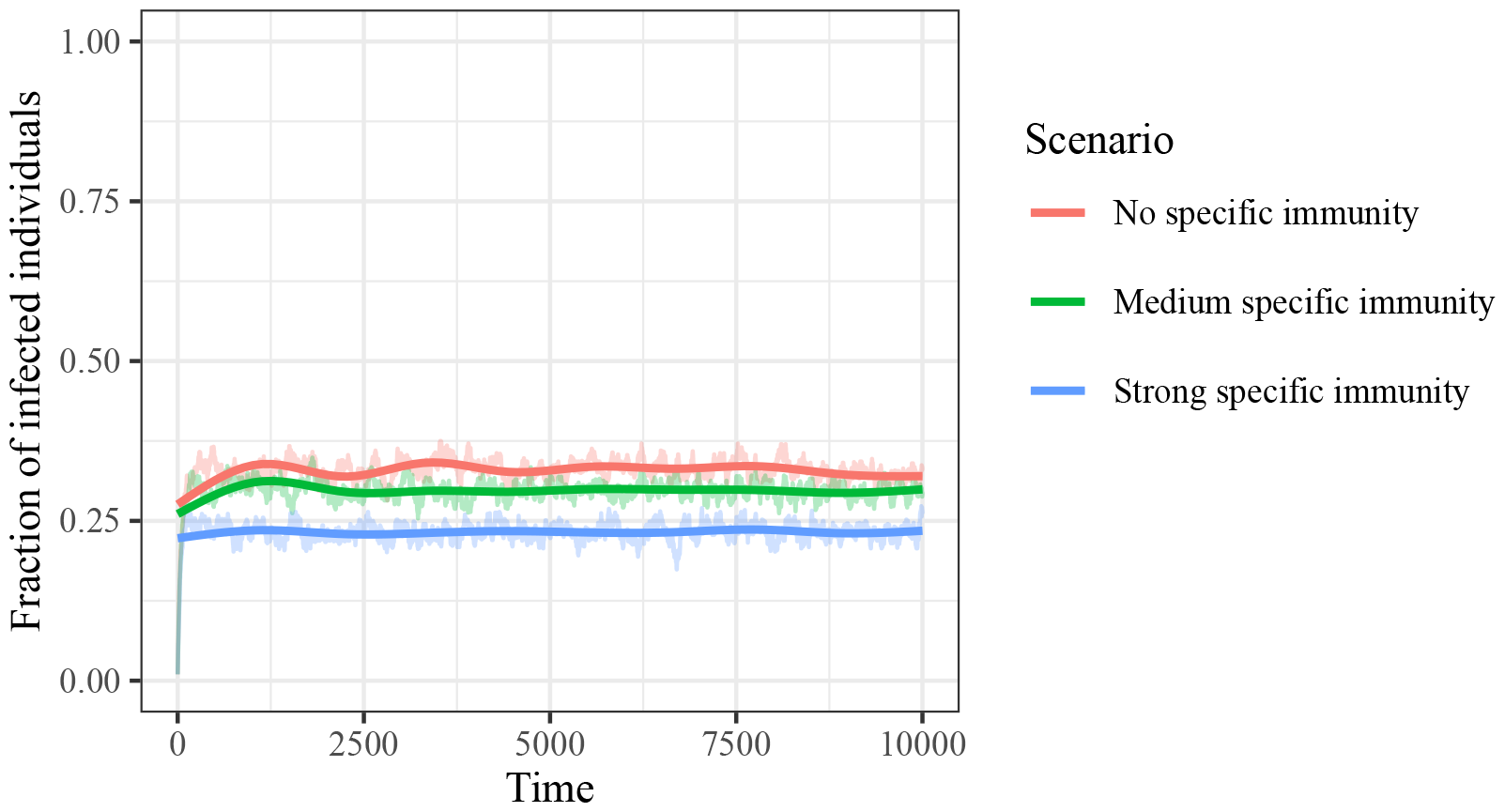
Dynamics in the compartments. The system oscillates around an infection level of about 25% in all three scenarios. The solid line is based on locally estimated scatter plot smoothing. The qualitative system dynamics are independent of the starting conditions, provided extinction of the disease does not occur (simulations not shown).

In order to illustrate the model dynamics, relation between the infection rate and the mean residence time in the two compartments is visualized in figure 7. For compartment **S**, we can observe that individuals with large values for *β*_*i*_ tend to have longer residence times. This is to be expected, as exposure to the infectious virus is less likely for small values of *β*_*i*_. The transition from **S** to **I** is identical for all three scenarios, so that there are no relevant differences between the three scenarios in this regard. In contrast, the mean residence time in **I** displays important differences between the three scenarios. In scenario ‘No specific immunity’, the duration of infections (= residence time in **I**) shows no dependence of the infection rate. For scenario “Medium specific immunity”, it can be seen that individuals with a higher infection rate (large *β*_*i*_) tend to have shorter infections. This relation is even more pronounced in the scenario “Strong specific immunity”.

**Figure 7.**
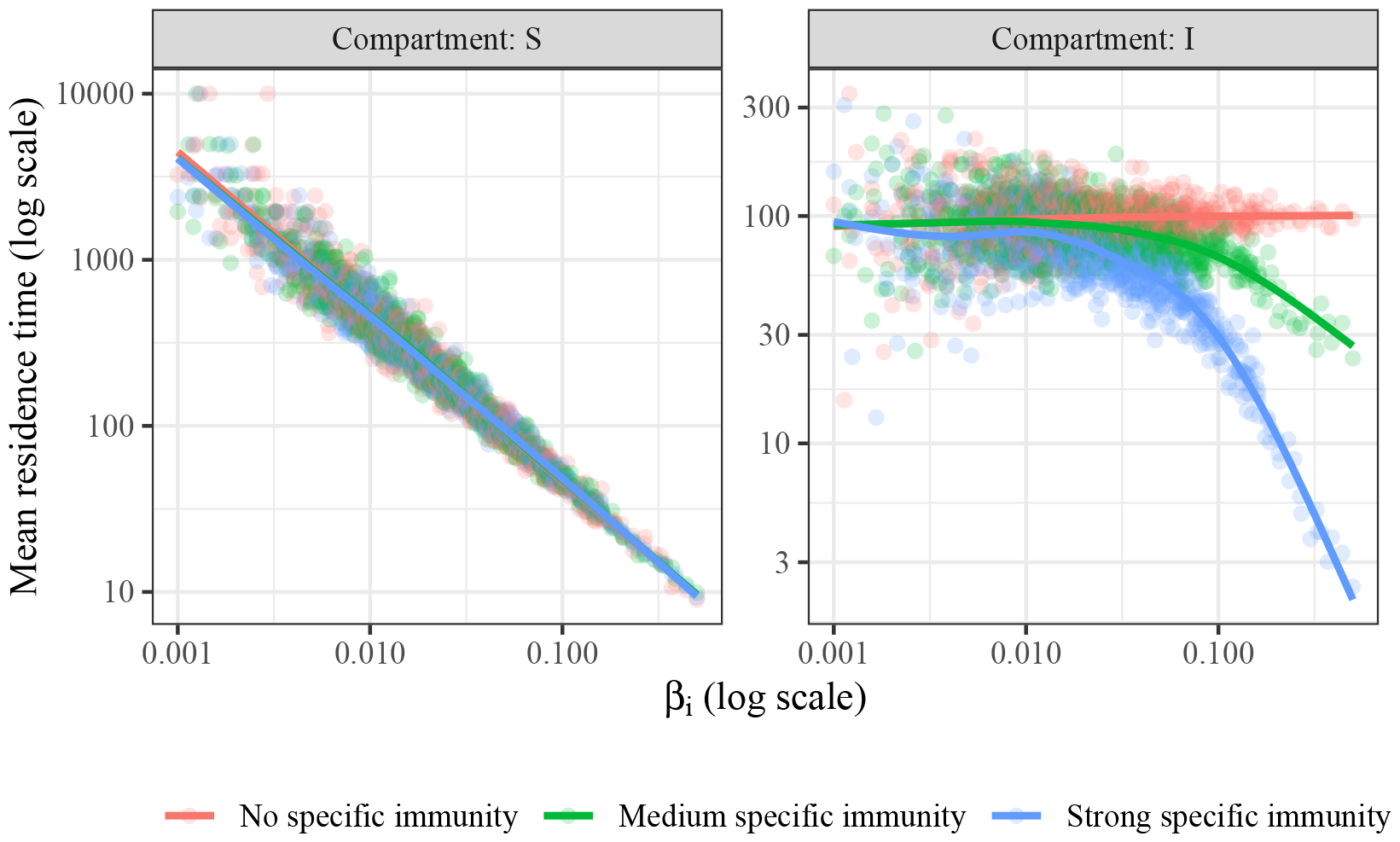
Mean residence times in the compartments **S** and **I**. For all individuals, the mean residence times in **S** and **I** have been calculated and plotted as individual dots on log scale versus the infection rate *β*_*i*_ on log scale. The solid lines are based on locally estimated scatter plot smoothing.

In figure 8, we show individual trajectories for the time interval from *t* = 9000 to *t* = 10000. It becomes clear that in all three scenarios, individuals with a small *β*_*i*_ are rarely infected and individuals with larger *β*_*i*_ have infections more frequently. In the first scenario (“No specific immunity”), the duration of infections is independent of *β*_*i*_. In the second scenario (“Medium specific immunity”), infections appear to be shorter for larger values of *β*_*i*_. In the third scenario (“Strong specific immunity”), this notion becomes more evident. In this scenario, the total time spent in state **I** first increases with increasing infection rate *β*_*i*_. For large values of *β*_*i*_, infections are frequent, however, tend to be short in duration.

**Figure 8.**
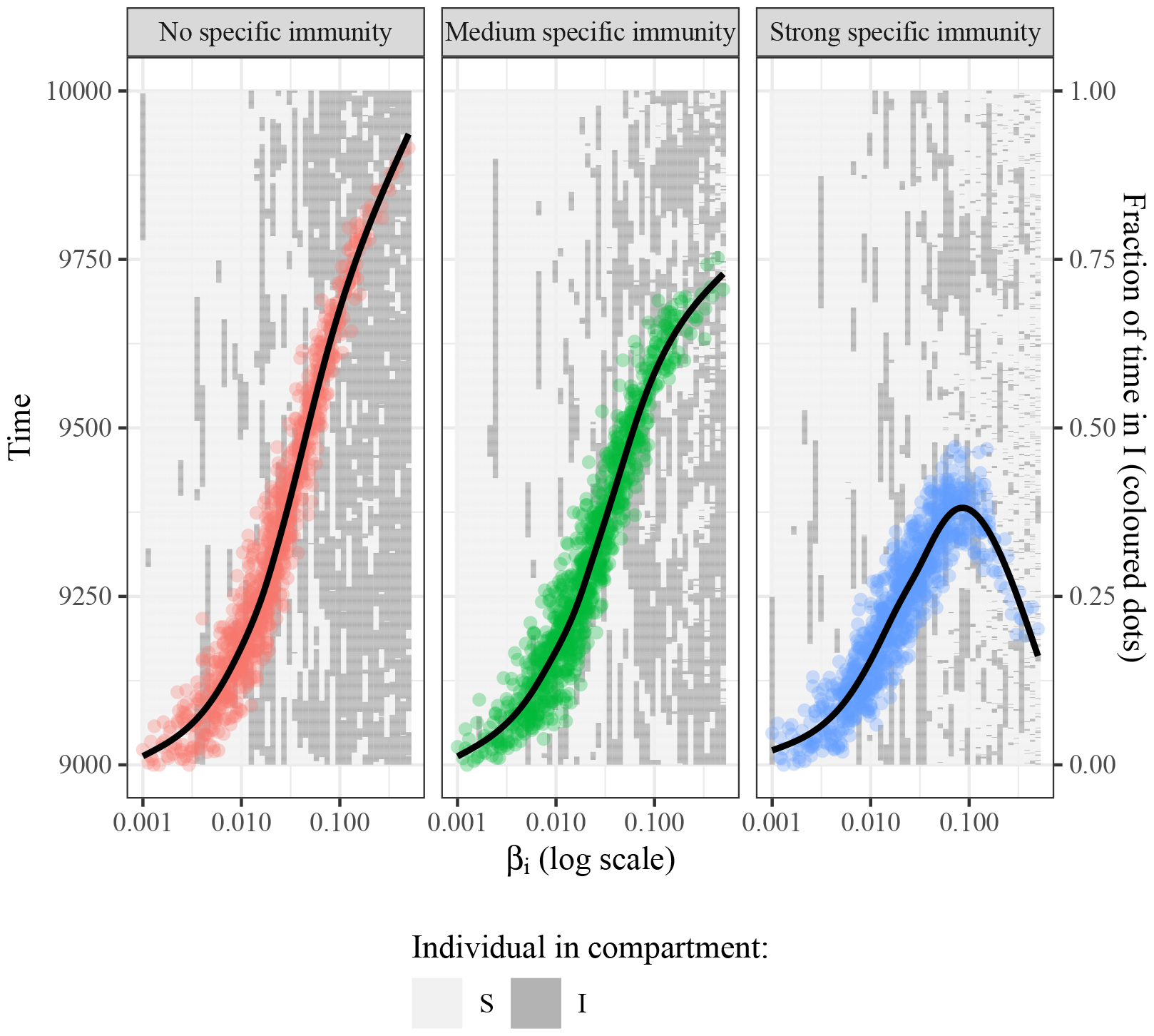
Trajectories of test individuals and mean disease burden. In this figure, time is plotted from bottom to top. Each column corresponds to a simulated test-individual with a particular value for *β*_*i*_. At the times marked dark gray, the individual is in **I**, at the times marked light gray in **S**. The colored dots and the black trend lines indicate the fraction of time spent in **I** with respect to the entire simulation time. The corresponding scale is on the right hand side of the figure.

The fraction of time spent in **I** in the simulation interval (*t* from 0 to 10000) is chosen as a proxy for mean disease burden per time. The fraction of time spent in **I** is the product of the number of infections and the mean residence time divided by the total time. In the scenarios “No specific immunity” and “Medium specific immunity”, there is a monotonous trend to increase with greater *β*_*i*_ (see trend line and colored dots in figure 8). However, in the scenario “Strong specific immunity”, the mean disease burden tends to decline once the infection rate *β*_*i*_ (and hence the contact rate) surpasses a certain value of *β*_*i*_. Hence, we confirm in our model analysis that also in populations that are heterogeneous with respect to their contact rates, an increased exposure frequency can lead to a decreased mean disease burden for appropriate parameter configurations, as already examined for the CC-ODE model.

## 4 DISCUSSION

The common cold causes considerable human distress and imposes substantial economic costs. It is widely assumed that adopting contact reduction, physical distancing and disinfection measures can reduce infections and that these measures are suited to reduce associated human suffering and economic burden. Although this may be true in many circumstances, it is not clear if this reasoning is universally valid. Interestingly, the relation between exposure frequency and mean disease burden has not been studied explicitly from a mathematical modeling perspective before.

In this paper, we present novel mathematical approaches that study the relation between pathogen exposure frequency and mean disease burden. We deliberately kept the presented approaches simplistic, yet they yield informative insights. Specifically, in this work we introduce an ordinary differential equation model derived from the SIRS model and an individual-based model derived from the SIS model. We apply the models to different theoretical scenarios in order to investigate this relationship.

In both modeling approaches, we demonstrate that for appropriate parameter constellations (i. e. properties of the virus and the population) an increased exposure may lead to a reduced mean disease burden. This can be explained by an efficient training of the immune system in the case of frequent infections. On the contrary, by reducing the number of infections, development of adequate immunity on an individual and population level may be hampered. Therefore, parameter configurations with high habituation rates arise, for which our models predict a reduced mean symptom burden with an increased exposure to a particular common cold virus, both on population and individual level. It is important to note that our models rely on the assumption that every exposure to the virus, even if it is not linked to a symptomatic infection (CC-ODE model) or associated with only a very short infection (CC-IB model) leads to full immunization. Further modeling analyses could be targeted at investigating the effect of relaxing this assumption.

For the novel CC-ODE model, the resulting properties and overall model behavior can be calculated analytically for given parameter configurations as demonstrated in section 3.1. While an absolute isolation of contagious individuals always leads to zero infections, this radical strategy is connected to great economic cost and personal restrictions. For rather harmless diseases such as common colds, this strategy is certainly not practicable. On the contrary, the better option could be to increase the contact rate in order to reduce the overall disease burden (if practically feasible). We can observe a decreasing mean disease burden with a larger overall contact rate after a certain threshold for most parameter configurations. The optimal strategy highly depends on this threshold and is probably different for each pathogen and considered population. For most people, the number of contacts of each individual is limited by external circumstances in practice such as occupation and lifestyle and can only be manipulated at a certain cost to the individual. This has to be evaluated carefully for the specific scenario. It should be noted here that the increase in contact is only ethically justifiable if the death rate is close to zero. In the case of common cold pathogens this should usually hold true. However, we definitely do not want to recommend increasing the contact rate for serious illnesses, even if this could lead to fewer infections.

The CC-IB model allows for studying the effects of heterogeneity, e. g. regarding contact behavior and / or development of immunity after exposure. In the current approach presented in section 3.2, we introduce heterogeneity exclusively with respect to the contact rate. For reasons of simplicity, the development of immunity and the resulting recovery dynamics are assumed to be identical for all individuals. A further level of heterogeneity with regard to the recovery dynamics could be introduced in the model, but that was not necessary for the investigation of the hypothesis examined in this paper. Furthermore, the individual-based approach adds stochastic aspects, both for infection and recovery. Analogous to the CC-ODE model, the results hint at the possibility that there might be constellations in which an increased exposure leads to a reduced mean disease burden with respect to specific viruses.

The parameters of the two models are related in a qualitative way. The infection rate *β* is described by infectivity *β*_1_ and the contact rate *β*_2_ in the CC-ODE model and by *β*_*i*_ in the CC-IB model. The specific immunogenicity is given by *α* and *a*, respectively. The parameter *γ* in the CC-ODE model and the parameter *c* in the CC-IB model correspond to the unspecific immunity. The time scale / duration of the immunity is modeled by the rate *δ* in the population-based approach and by *d* in the individual-based approach.

In conclusion, the CC-ODE model is more targeted at global decision-making issues, that might arise in public health politics. This approach is more suited to derive public health strategies to reduce the average disease burden. On the contrary, if the concern is to give specific advice to patients, the CC-IB model can give more insightful recommendations. Furthermore, the individual-based model allows to study the dynamics of populations with heterogeneous contact rates, which is not possible with the ordinary differential equation approach presented in this paper. However, both models capture the notion that an increased exposure frequency leads to a reduced disease burden for certain parameter constellations. The models are rather simplistic in design and do not implement all possible facets of interaction between individuals and common cold viruses.

Up to this point, we have chosen illustrative model parameters for some generic common cold virus. In reality, there is a large diversity of the circulating virus. Furthermore, the ensemble of circulating viruses is not static but underlies constant development, in particular as a consequence of mutations of existing viruses and appearance of new viruses and virus variants from other species. The immunological memory within individuals and also within a population is shaped by the exposure to this multitude of different viruses. Furthermore, cross-immunity between different viruses and strains of viruses causing common colds may have a significant impact on disease patterns. These levels of complexity are currently not included in the presented modeling approaches. However, it is possible to make use of our models as a starting point and extend them to include multiple virus strains and interactions, for instance.

In case of the CC-ODE model, with knowledge of the average time of infection an estimate for the recovery rate *γ* can be provided, while the average time between infections determines the immunity loss rate *δ*. The population numbers for the three different states over time and the basic reproduction number can help to determine the other parameters. Thereby, the parameter estimation also highly depends on the available data and the framework and one also has to consider the time scale and population size. In this way, a realistic scenario for an existing pathogen can be evaluated by fitting the model parameters. As a consequence, the model predictions could be challenged, in particular the hypothesis that a higher frequency of exposure can lead to a lower mean disease burden.

In the case of common cold pathogens that do not lead to protective immunity (e. g. parainfluenza viruses, metapneumoviruses), there is insufficient evidence and data to answer the following hypothetical question: Would a person exposed to a specific common cold pathogen (e. g. a particular rhinovirus) at high frequency (e. g. once a day / hourly) experience persistent corresponding symptoms? Furthermore, the impact of cross-reactivity between different virus serotypes is also not well-studied. For non-common-cold infectious agents, there is a small number of studies systematically investigating intensity of symptoms upon repeated exposure (Dittmer et al., 1995; Hattakam et al., 2021; De Angelis et al., 2021; Frumento et al., 2022).

During the COVID-19 pandemic, many regions were subject to long-lasting, extensive contact restrictions, particularly in the winters of 2020/2021 and 2021/2022. As a consequence of the reduced contacts, the incidence of common colds has dropped during this time. This is presumably linked to a general decline in the population’s immunity to cold viruses. In the winter 2022/2023, an increased number of sick days due to common colds was observed in the communities where the contact rates had been lower in the preceding years. This can be explained in a straight-forward manner with the presented models.

In order to test the results derived in this analysis, different experimental strategies are conceivable. First, in an animal study, animals could be repeatedly infected with a typical common cold virus (e. g. a rhinovirus) in a controlled setting, systematically varying the time interval between subsequent infections and recording the intensity of the disease symptoms. Appropriate animal models have been described in the literature (Yin and Lomax, 1986). Second, humans could be repeatedly infected with different time intervals with a mild common cold virus. In an investigation studying the relation between sleep duration and intensity of symptoms of a common cold, the authors chose such an approach (Prather et al., 2015). From our point of view, however, it is questionable whether such a procedure is ethically justifiable for our research question for both animal and human studies, as even generally mild pathogens can lead to more severe courses of disease in rare cases. A third conceivable approach is to collect observational data from groups with different exposure rates to common cold viruses via questionnaires. The exposure frequency most likely varies considerably between different professions. An attempt could be made to quantify the exposure and symptom burden of common cold infections in a prospective longitudinal study, ideally including participants with a wide range of different contact behaviors. A fourth approach might be related to a more comprehensive surveillance of common cold pathogens by federal agencies, as is already being done (e. g. weekly reports by the Robert-Koch-Institut in Germany - https://influenza.rki.de/Wochenberichte.aspx).

A general increase of contacts in order to stimulate training of the specific immunological defense seems neither feasible nor desirable. Foremost, there are not only mild pathogens in circulation. Viruses such as SARS-CoV-2 and influenza lead to severe, often life-threatening and in many cases lethal infections. The presence of dangerous infectious agents is prohibitive of calling for a general increase of contacts. Nonetheless, the considerations raised in this article raise the question of whether the repertoire of infectious agents to which the immune system is exposed could be specifically targeted in a novel way. In certain environments such as childcare facilities or schools, for example, constellations are conceivable in which avoiding contact (as oftentimes recommended in the cold seasons) could lead to a worsening of the disease burden due to common colds. Loosely speaking, an infection with a benign pathogen that can be successfully averted today might be the common cold of tomorrow with more severe symptoms due to a prospective decreased specific immunity.

Another measure to increase specific immunity against common cold pathogens and thereby reduce the associated disease burden are vaccination approaches (Simancas-Racines et al., 2017). However, such approaches have turned out to be challenging due to the multitude and ongoing evolution of the causative agents. To date, there are no such vaccines available that are recommended in the relevant guidelines. As common cold infections are usually mild, research in this field is limited and no groundbreaking innovations are to be expected in the near future.

In conclusion, a more detailed characterization of disease dynamics of the common cold seems worthwhile. To this end, more detailed observational and experimental data are required in order to facilitate more specific mathematical modeling approaches. More sophisticated mathematical models fitted to more precise data potentially allow to derive recommendations suited to decrease the disease burden associated with the common cold.

## Data Availability

The source code of all analyses performed in this study are available upon reasonable request to the authors.

## REFERENCES

Arruda, E., Pitkäranta, A., Witek Jr, T. J., Doyle, C. A., and Hayden, F. G. (1997). Frequency and natural history of rhinovirus infections in adults during autumn. Journal of clinical microbiology 35, 2864–2868

Bramley, T. J., Lerner, D., and Sarnes, M. (2002). Productivity losses related to the common cold. Journal of occupational and environmental medicine, 822–829

De Angelis, M. L., Francescangeli, F., Rossi, R., Giuliani, A., De Maria, R., and Zeuner, A. (2021). Repeated exposure to subinfectious doses of sars-cov-2 may promote t cell immunity and protection against severe covid-19. Viruses 13, 961

DeAngelis, D. L. and Grimm, V. (2014). Individual-based models in ecology after four decades. F1000prime reports 6

Dittmer, U., Stahl-Hennig, C., Coulibaly, C., Nisslein, T., Lüke, W., Fuchs, D., et al. (1995). Repeated exposure of rhesus macaques to low doses of simian immunodeficiency virus (siv) did not protect them against the consequences of a high-dose siv challenge. Journal of general virology 76, 1307–1315

Frumento, N., Figueroa, A., Wang, T., Zahid, M. N., Wang, S., Massaccesi, G., et al. (2022). Repeated exposure to heterologous hepatitis c viruses associates with enhanced neutralizing antibody breadth and potency. The Journal of Clinical Investigation 132

Glanville, N., Mclean, G. R., Guy, B., Lecouturier, V., Berry, C., Girerd, Y., et al. (2013). Cross-serotype immunity induced by immunization with a conserved rhinovirus capsid protein. PLoS pathogens 9, e1003669

Hattakam, S., Elong Ngono, A., McCauley, M., Shresta, S., and Yamabhai, M. (2021). Repeated exposure to dengue virus elicits robust cross neutralizing antibodies against zika virus in residents of northeastern thailand. Scientific Reports 11, 9634

Heikkinen, T. and Järvinen, A. (2003). The common cold. The Lancet 361, 51–59

Kermack, W. O. and McKendrick, A. G. (1932). Contributions to the mathematical theory of epidemics. ii.—the problem of endemicity. Proceedings of the Royal Society of London. Series A, containing papers of a mathematical and physical character 138, 55–83

Murphy, K., Weaver, C., Berg, L., and Barton, G. (2022). Janeway’s Immunobiology (W W Norton & Co Inc), 10 edn.

Passioti, M., Maggina, P., Megremis, S., and Papadopoulos, N. G. (2014). The common cold: potential for future prevention or cure. Current allergy and asthma reports 14, 1–11

Prather, A. A., Janicki-Deverts, D., Hall, M. H., and Cohen, S. (2015). Behaviorally assessed sleep and susceptibility to the common cold. Sleep 38, 1353–1359

Simancas-Racines, D., Franco, J. V., Guerra, C. V., Felix, M. L., Hidalgo, R., and Martinez-Zapata, M. J. (2017). Vaccines for the common cold. Cochrane Database of Systematic Reviews

Thomas, M. and Bomar, P. A. (2022). Upper Respiratory Tract Infection (StatPearls Publishing, Treasure Island (FL))

Turner, R. B. (2015). The common cold. Mandell, Douglas, and Bennett’s principles and practice of infectious diseases, 748

Weiss, G. H. and Dishon, M. (1971). On the asymptotic behavior of the stochastic and deterministic models of an epidemic. Mathematical Biosciences 11, 261–265

Yin, F. H. and Lomax, N. B. (1986). Establishment of a mouse model for human rhinovirus infection. Journal of general virology 67, 2335–2340

