## Supplementary material for "Impact of exposure frequency on disease burden of the common cold - a mathematical modeling perspective"

medRχiv
THE PREPRINT SERVER FOR HEALTH SCIENCES

### Supplementary Material

#### 1 STABILITY OF NON-TRIVIAL STEADY STATE OF CC-ODE MODEL

The stability condition for the non-trivial steady state is computed with the help of the eigenvalues of the Jacobian at the non-trivial steady state:

$$\lambda_{1/2} = \frac{-\delta(\beta_1\beta_2+\delta)}{(2\alpha\beta_2+\gamma+\delta)} \pm \frac{\delta^{0.5}}{(2\alpha\beta_2+\gamma+\delta)} \cdot \Big(4\alpha^3\beta_2^3 - 4\alpha^2\beta_1\beta_2^3 + 12\alpha^2\gamma\beta_2^2 + \tag{S1}$$

$$8\alpha^2\delta\beta_2^2 - 8\alpha\beta_1\gamma\beta_2^2 - 8\alpha\beta_1\delta\beta_2^2 + 12\alpha\gamma^2\beta_2 + 16\alpha\gamma\delta\beta_2 + 4\alpha\delta^2\beta_2 + \beta_1^2\delta\beta_2^2 - \tag{S2}$$

$$4\beta_1\gamma^2\beta_2 - 8\beta_1\gamma\delta\beta_2 - 2\beta_1\delta^2\beta_2 + 4\gamma^3 + 8\gamma^2\delta + 4\gamma\delta^2 + \delta^3\Big)^{0.5}, \tag{S3}$$

$$\lambda_3 = 0. \tag{S4}$$

For the choice of $\delta = 0.1$, we obtain the following behaviour depending on $\alpha$, $\beta_1$, $\beta_2$ and $\gamma$.

Generally speaking, the non-trivial steady state is stable for low values of $\alpha$ and $\gamma$ and high values of $\beta_1$ and $\beta_2$. The latter parameters represent the rate of spread of the disease and the higher they are, the more likely it will be for the disease to remain present in the population for a longer time period. Hence, the

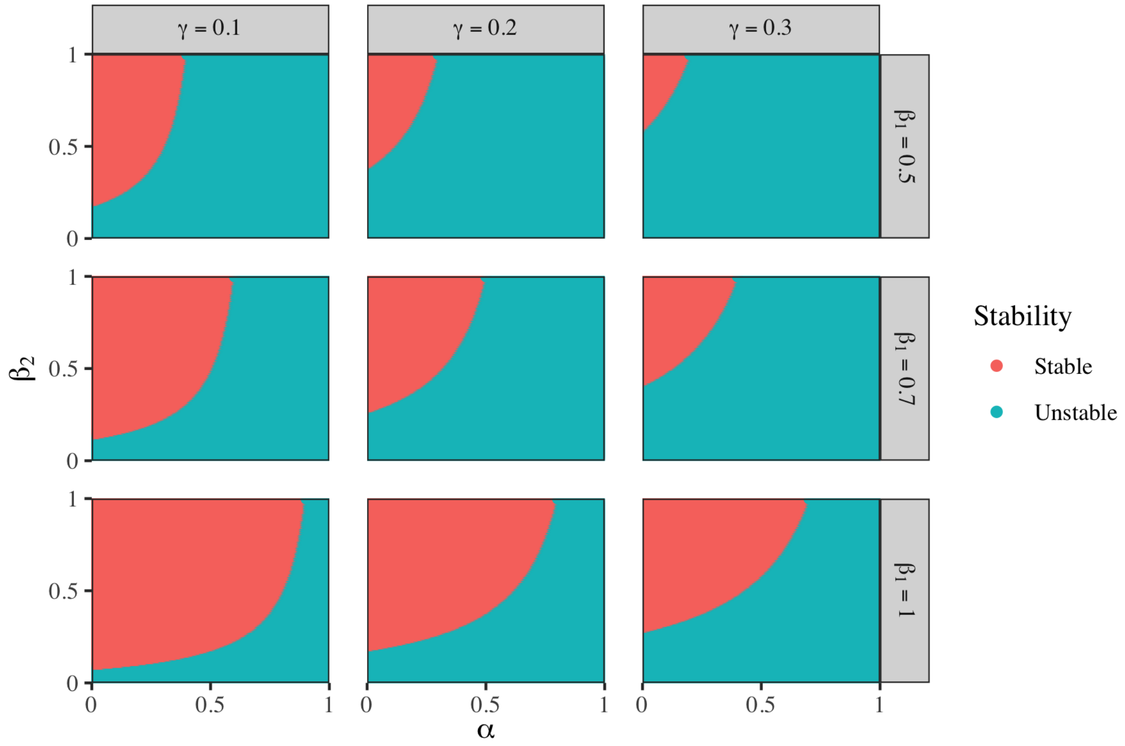


**Figure S1.** Stability of non-trivial steady state for the parameters $\beta_1 \in \{0.5, 0.7, 1.0\}$, $\gamma \in \{0.1, 0.2, 0.3\}$ and $\delta = 0.1$

non-trivial state that includes a constant proportion of infected individuals will be stable and there will always be infections. In contrast, the parameters $\alpha$ and $\gamma$ generate the flux towards the non-infected state $\mathbf{R}$ and finally to $\mathbf{S}$, if $\delta > 0$.
